# Adaptation and validation of a scale to evaluate the quality of virtual courses developed for medical students in Peru during the COVID-19 pandemic

**DOI:** 10.1101/2022.01.08.22268928

**Authors:** Claudio Intimayta-Escalante, Rubí Plasencia-Dueñas, Kevin Flores-Lovon, Janeth N. Nuñez-Lupaca, Mario Chavez-Hermosilla, Ronald Castillo-Blanco

## Abstract

**Background:** During the COVID-19 pandemic, medical education migrated to digital environments, without clear guidelines for virtual courses or evaluations of how these courses have been developed.

**Objective:** To adapt and validate a scale to evaluate the quality of virtual courses developed for human medicine students in Peru.

**Methods:** Cross-sectional study that adapted a scale to assess the quality of virtual courses to the context of Peruvian medical students during the COVID-19 pandemic, using the Delphi methodology and pilot tests for a rigorous evaluation of the items, resulting in a scale of 30 items that were described with summary statistics. In addition to the exploratory factor analysis (EFA) with Oblimin rotation, together with the adequacy and sample fit with Bartlett test and Kaiser-Meyer-Olkin (KMO), while the internal consistency was estimated with the alpha coefficient.

**Results:** A total of 297 medical students in Peru were surveyed. The descriptive statistics for the items showed a normal distribution, while the Bartlett test showed no inadequacy (X2=6134.34, p<0.01) and with the KMO test an overall value greater than 0.92 was found, therefore an AFE was performed where five factors were identified (General Quality and Didactic Methodology, Design and Navigation of the Virtual Platform, Multimedia Resources, Academic Materials) with 30 items. In the internal consistency, an alpha coefficient greater than 0.85 was estimated for the factors evaluated.

**Conclusions:** The adapted scale of 30 items grouped into five factors or domains, show adequate evidence of validity and reliability to be used in the evaluation of the quality of virtual courses developed for Peruvian human medicine students during the context of the COVID-19 pandemic.

## Introduction

In recent years, virtual courses have been progressively integrated into university education (1). However, the COVID-19 pandemic caused drastic changes in education, forcing a complete migration to digital environments (2,3). This had further repercussions on the university education of medical students who had an interruption in their classes and face-to-face practices in hospital or community centers (4,5) due to the social distancing norms and the saturation of health centers due to the large number of patients with COVID-19 (6).

The rapid impact of the pandemic in Peru emphasized the importance of improving the formation of future health professionals (7). Therefore, it was necessary for universities, faculties and schools with medical students to adapt to virtual environments in an effective and practical way to continue with the training process (8-10). However, many institutions showed limitations and were not prepared for this sudden change, with an absence of virtual platforms together with scarce training and support for teachers who were mostly health personnel (11,12), all this evidences the need for an improvement of the virtual courses that were carried out without clear directives or focused on the needs of the different university careers such as those of health sciences (13).

In this context, more than a year after the pandemic despite the instruments and recommendations that currently exists (14), many institutions responsible for the formation of medical students in Peru have yet to carry out critical evaluations of how virtual courses have been developed to improve them. Therefore, the aim of the present study was to culturally adapt and validate a scale to evaluate the quality of virtual courses developed for human medical students in Peru during the COVID-19 pandemic.

## Methods

### Survey population

Cross-sectional study, where Peruvian human medicine students enrolled in undergraduate courses corresponding to the academic year 2020 were surveyed. We excluded those respondents whose medical schools did not use virtual classrooms for the courses or did not answer all the questions of the scale or answered the survey twice, considering in these cases only the first answer. For an adequate evaluation and applicability of the instrument, students from the majority of human medical schools in Peru were surveyed by non-probabilistic convenience sampling.

### Cultural adaptation

The adaptation was carried out with the Delphi methodology using a 36-item scale developed by Santoveña S. et al. to evaluate the quality of the complementary virtual courses developed for the Social Education career at the Universidad Nacional de Educación a Distancia (UNED) in Spain, considering three dimensions: quality of the environment, quality of the didactic methodology used and technical quality (15). The members of the committee for the adaptation of the instrument were five researchers with experience in the area: two with knowledge of psychometric analysis methodology, two with experience in medical education research and one with teaching experience in human medical schools. In this way, the appropriateness of the adapted version for medical students and the content-based validity of the items were evaluated with Aiken’s V indicator.

### Pilot study

After the evaluation by the members of the committee for the adaptation of the instrument, two pilot studies were carried out to evaluate the final adapted version with 36 items in students from 43 human medical schools in Peru, seeking to evaluate the understandability of the items and the applicability in the different human medical schools at the national level. After four weeks of the first pilot study, the scale was modified to 32 items considering the comments of the students and the recommendations of Furr R. for the development of measurement instruments in health (16), so a new pilot study was conducted with the final version of the scale and other sections of the survey focused on general characteristics such as age, sex, academic year and type of university management, were also evaluated by the participants of the pilot test at this stage.

### Data collection

The data were collected with a self-applied online survey (https://is.gd/Calidad20) developed in REDCap v.8.1.8 software (Research Electronic Data Capture version 8.1.8) disseminated since july 2021 through social networks (Facebook, Whatsapp and Instagram) with drawings among respondents to encourage their participation and dissemination. The survey consisted of multiple specific sections: one for general data and another for the scale statements. For scale items that shared a similar opening sentence, a common heading was placed in that section of the survey for the items as appropriate **(Table 1)**. The response options were in Likert scale format that evaluates the participant’s degree of agreement with the statements of the items (strongly disagree, disagree, neither agree nor disagree, agree or strongly agree), scoring from one to five each of the response options, considering the order mentioned above.

**Table 1.** Structure and descriptive statistics of the scale to evaluate the quality of the virtual courses developed for Peruvian students of human medicine during the COVID-19 pandemic.

| No. | Detailed item text | *M* | *SD* | *g*<sub>1</sub> | *g*<sub>2</sub> |
| --- | --- | --- | --- | --- | --- |
| **The virtual courses...** |  |  |  |  |  |
| 2 | ...had a quality proportional to the investment. | 3.10 | 0.97 | - 0.45 | - 0.22 |
| 3 | ...had varied and interesting activities. | 2.96 | 0.90 | - 0.24 | - 0.09 |
| 4 | ...promoted the understanding and reasoning of the subjects. | 3.25 | 0.92 | - 0.48 | - 0.19 |
| 5 | ...were organized in the development of their contents (developing an introduction, the objectives and the different topics of each chapter). | 3.37 | 0.96 | - 0.60 | 0.00 |
| 6 | ...had comprehensive and sufficient content for learning the topics. | 3.20 | 0.89 | - 0.34 | - 0.20 |
| 7 | ...addressed the contents in a clear way. | 3.35 | 0.88 | - 0.44 | 0.15 |
| **Teachers in virtual courses...** |  |  |  |  |  |
| 8 | ...used a teaching methodology that encouraged reflection and/or questions about the topics. | 3.15 | 0.91 | - 0.38 | - 0.42 |
| 10 | ...developed assessments appropriate to the content developed. | 3.32 | 0.95 | - 0.70 | - 0.17 |
| 11 | ...provided adequate feedback for the assignments or evaluations developed. | 2.93 | 1.01 | - 0.36 | - 0.66 |
| **Digital media used for the courses...** |  |  |  |  |  |
| 12 | ...facilitated the teaching-learning process (they made it possible to organize work groups, create forums, provide feedback, and others). | 3.42 | 0.88 | - 0.57 | 0.15 |
| 13 | ...allowed the development of new ideas or interpretations that favored the discussion of the topics. | 3.20 | 0.93 | - 0.33 | - 0.32 |
| **The virtual classroom...** |  |  |  |  |  |
| 14 | ...had a nice and balanced format (adequate text size and image quality). | 3.44 | 0.95 | - 0.68 | 0.10 |
| 15 | ...had an adequate organization (hierarchy of topics and index of contents). | 3.51 | 0.92 | - 0.75 | 0.24 |
| 16 | ...had well-designed and functional headers, titles and buttons. | 3.54 | 0.93 | - 0.64 | 0.04 |
| 17 | ...had pages of adequate length (short texts, hyperlinks, quick access to main menu or similar). | 3.40 | 0.92 | - 0.62 | - 0.07 |
| **Tools in the virtual classroom...** |  |  |  |  |  |
| 18 | ...had pages of adequate length (short texts, hyperlinks, quick access to main menu or similar). | 3.39 | 0.86 | - 0.61 | - 0.09 |
| 19 | ...allowed a good navigation among the contents. | 3.42 | 0.88 | - 0.52 | 0.08 |
| 20 | ...facilitated interaction between users (students and/or teachers). | 3.23 | 0.91 | - 0.37 | - 0.53 |
| 21 | ...helped to better study the contents developed in the course. | 3.17 | 0.85 | - 0.16 | - 0.29 |
| 22 | ...helped in the evaluation of the courses (allowed the development of exams, visualization of grades, etc.). | 3.48 | 0.95 | - 0.62 | 0.10 |
| **The Multimedia resources (videos, simulators, etc.) used in the courses...** |  |  |  |  |  |
| 23 | ...were characterized by a quality design integrated to the structure of the virtual classroom. | 3.10 | 0.97 | - 0.36 | - 0.50 |
| 24 | ...had a general description, transcription and content labels. | 3.00 | 0.98 | - 0.27 | - 0.72 |
| 25 | ...were varied and integrated to the topics developed. | 3.11 | 0.96 | - 0.47 | - 0.47 |
| 26 | ...were useful for the assigned tasks. | 3.20 | 0.98 | - 0.53 | - 0.37 |
| **Videoconferences...** |  |  |  |  |  |
| 27 | ...were scheduled well in advance. | 3.53 | 0.98 | - 0.63 | - 0.22 |
| 28 | ...were posted in the virtual classroom for later review. | 3.27 | 1.14 | - 0.38 | - 0.78 |
| 29 | ...were conducted at an adequate pace. | 3.23 | 1.02 | - 0.38 | - 0.46 |
| **Academic materials (books, articles, or similar) used in the courses...** |  |  |  |  |  |
| 30 | ...had understandable contents. | 3.56 | 0.81 | - 1.15 | 1.39 |
| 31 | ...contained updated information. | 3.52 | 0.89 | - 0.62 | 0.13 |
| 32 | ...were in line with the learning objectives. | 3.62 | 0.81 | - 0.84 | 0.92 |

**No.:** item number, **M:** mean, **SD:** standard deviation, **g**<sub>1</sub>: skewness, **g**<sub>2</sub>: kurtosis.
**Note:** To obtain the Spanish version of scale, please write to the corresponding author.

### Statistical analysis

The statistical analysis was developed in R Studio software version 3.4 with the packages “psych”, “lavaan”, “semTools”, “semPlot” and “GPArotation”, using absolute and relative frequency measures to describe the general characteristics of the respondents, using measures of central tendency (mean) and dispersion (standard deviation) for the numerical variables. In the descriptive analysis, skewness and kurtosis were estimated, while in the psychometric analysis to assess validity based on internal structure, an exploratory factor analysis was performed using the unweighted least residuals estimator (ULS) and Oblimin’s oblique rotation to determine the number of factors or domains. All this considering the polychoric correlation between items and the communality of each item. In addition, the overall sample adequacy was measured by means of Bartlett test of sphericity and the Kaiser-Meyer-Olkin test (KMO), taking as a correct adjustment of the average sample and per item when the values were higher than 0.80 (17). As for the fit indexes, the residual root mean square root (RMSR) was used. For the reliability analysis, internal consistency was approximated by calculating the alpha coefficient.

### Ethical aspects

The study did not suppose a risk for the respondents or researchers, since it was conducted virtually, respecting the rules of physical distancing and ensuring confidentiality, as well as anonymity, without informing the name of the universities where the respondents were studying and codifying the identification data that were used to verify that the respondents are medical students. In addition, the study protocol was submitted and reviewed by the research ethics committee of the Hospital Nacional Almanzor Aguinaga Asenjo of Peru, obtaining the ethical approval for the study with the resolution NTI-75502021030, this was obtained before of starting the pilot study and data collection.

## Results

A total of 337 Peruvian human medicine students were surveyed; however, after applying the exclusion criteria, there were 297 medical student surveys, with an average of seven students for each of the 41 human medicine schools in Peru. More than half of the participants were female (53%), with an overall average age of 22.50 years (SD=3.43). Regarding distribution by academic year, 24.58% were third year (n=73), 22.55% were fourth year (n=64), 19.53% were fifth year (n=58), 15.82% were second year (n=47), 15.49% were first year (n=46) and 3.03% were sixth year (n=9). The majority of respondents (69.02%) studied in public universities as opposed to 92 students from private universities.

After the evaluation by the five committee members for the cultural adaptation in two rounds, a 0.85 of agreement was identified between members, with a range between 0.80 and 0.96 scores given by the committee members.

In the descriptive analysis of the scale, it was found that the means of the values of the items were between 2.93 and 3.62 while the standard deviation was in the range of 0.81 and 1.14, in addition, the skewness values were between -1.15 and -0.15 and the kurtosis between -0.78 and 1.39, showing values suitable for a normal distribution **(Table 1)**.

Regarding the psychometric indicators of the scale composed of 32 items, two items were identified with low factor loads and a low correlation with respect to the rest of the items, which led to their exclusion, being item 1 (“Virtual courses had a syllabus or syllabus before they started”) with a factor load of 0. 29 and item 9 (“The teachers in the virtual courses gave a reasonable number of assignments”), which despite a factor load of 0.43, showed a correlation with other items of less than 0.42.

In the third round, after the pilot studies, the evaluation of content-based validity with the committee members addressed the criteria of relevance, representativeness and clarity for the 30 items of the scale, an Aiken’s V value of 0.86 was obtained, with a range between 0.72 and 0.92 of score given by the committee members.

The evaluation with the KMO method identified an overall value of 0.95. On the other hand, with Bartlett test of sphericity, no inadequacy was detected (x^2^=6134.34, p<0.01), demonstrating adequate conditions to perform the exploratory factor analysis. Identifying five domains or factors for the scale, with the minimum residual estimator of the items and the Oblimin rotation, in addition, the factor loadings of the items considered for the final model were between 0.45 and 0.91, together with communality values between 0.31 and 0.93 **(Table 2)**.

**Table 2.** Results of the analysis of the factors of the scale to evaluate the quality of the virtual courses developed for Peruvian students of human medicine during the COVID-19 pandemic.

| No. of item | Specific value of factor loadings |  |  |  |  | h<sup>2</sup> |
| --- | --- | --- | --- | --- | --- | --- |
|  | Factor 1 | Factor 2 | Factor 3 | Factor 4 | Factor 5 |  |
| 2 | **0.50** | 0.01 | 0.11 | - 0.12 | 0.06 | 0.31 |
| 3 | **0.70** | 0.04 | 0.18 | - 0.13 | 0.02 | 0.62 |
| 4 | **0.75** | 0.00 | 0.07 | 0.01 | 0.04 | 0.67 |
| 5 | **0.70** | 0.02 | 0.05 | 0.14 | - 0.01 | 0.67 |
| 6 | **0.63** | 0.03 | 0.06 | 0.14 | - 0.06 | 0.55 |
| 7 | **0.78** | 0.03 | - 0.03 | 0.14 | - 0.04 | 0.70 |
| 8 | **0.69** | - 0.05 | 0.17 | - 0.03 | 0.09 | 0.68 |
| 10 | **0.62** | 0.19 | - 0.03 | 0.04 | - 0.03 | 0.54 |
| 11 | **0.62** | - 0.11 | 0.05 | 0.15 | 0.13 | 0.57 |
| 12 | **0.45** | 0.17 | 0.12 | - 0.13 | 0.24 | 0.56 |
| 13 | **0.55** | 0.06 | 0.21 | - 0.14 | 0.18 | 0.63 |
| 14 | - 0.04 | **0.71** | 0.11 | 0.01 | 0.16 | 0.75 |
| 15 | 0.04 | **0.83** | 0.08 | 0.07 | - 0.08 | 0.78 |
| 16 | 0.06 | **0.84** | - 0.01 | 0.01 | 0.04 | 0.82 |
| 17 | 0.12 | **0.66** | 0.02 | - 0.02 | 0.17 | 0.74 |
| 18 | 0.05 | 0.07 | 0.00 | 0.19 | **0.67** | 0.71 |
| 19 | - 0.14 | 0.32 | 0.03 | 0.12 | **0.67** | 0.81 |
| 20 | 0.20 | - 0.07 | 0.04 | 0.02 | **0.75** | 0.75 |
| 21 | 0.11 | 0.10 | 0.23 | - 0.07 | **0.59** | 0.72 |
| 22 | 0.04 | 0.10 | 0.07 | 0.10 | **0.56** | 0.56 |
| 23 | 0.04 | 0.00 | **0.77** | 0.04 | 0.07 | 0.75 |
| 24 | - 0.01 | - 0.03 | **0.91** | 0.01 | 0.07 | 0.86 |
| 25 | 0.06 | 0.10 | **0.80** | 0.03 | - 0.03 | 0.79 |
| 26 | 0.03 | 0.08 | **0.79** | 0.12 | - 0.08 | 0.75 |
| 27 | **0.54** | 0.11 | - 0.12 | 0.14 | 0.10 | 0.46 |
| 28 | **0.53** | 0.18 | - 0.10 | 0.05 | - 0.07 | 0.32 |
| 29 | **0.67** | 0.08 | - 0.11 | 0.10 | 0.08 | 0.56 |
| 30 | 0.19 | 0.25 | - 0.01 | **0.52** | 0.03 | 0.68 |
| 31 | 0.05 | - 0.02 | 0.15 | **0.70** | 0.09 | 0.70 |
| 32 | 0.08 | 0.03 | 0.10 | **0.83** | 0.07 | 0.93 |

| No. of factor | Specific value in the correlation | | | | | $\alpha$ |
| --- | --- | --- | --- | --- | --- | --- |
|  | Factor 1 | Factor 2 | Factor 3 | Factor 4 | Factor 5 |  |
| Factor 1 | - |  |  |  |  | 0.92 |
| Factor 2 | 0.53 | - |  |  |  | 0.90 |
| Factor 3 | 0.65 | 0.51 | - |  |  | 0.91 |
| Factor 4 | 0.53 | 0.49 | 0.41 | - |  | 0.86 |
| Factor 5 | 0.52 | 0.62 | 0.57 | 0.37 | - | 0.88 |

No.: number, h<sup>2</sup>: community, $\alpha$ : alpha coefficient.
\*The values of factor loadings highlighted are those considered for the final model.

Regarding the factors identified for the 30 items of the scale, it was found a proportion of variance explained in the first factor (“General Quality and Didactic Methodology”) of 0.35, in the second factor (“Design of the Virtual Platform”) of 0.18, in the third factor (“Multimedia Resources”) of 0.19, in the fourth factor (“Academic Materials”) of 0.12 and in the fifth factor (“Navigation in the Virtual Platform”) of 0.17. 18, in the third factor (“Multimedia Resources”) of 0.19, in the fourth factor (“Academic Materials”) of 0.12 and in the fifth factor (“Navigation in the Virtual Platform”) of 0.17, while the correlations between factors were identified with values between 0.37 and 0.65. Likewise, in the evaluation of the internal consistency of the factors, alpha coefficient values greater than 0.85 were found **(Table 2)**. In addition, in the fit indices, a value of 0.04 was estimated for the RMSR.

## Discussion

The adapted version of the scale to evaluate the quality of virtual courses for Peruvian medical students developed in the context of COVID-19 shows adequate indicators for its use in critical evaluations of these courses, which will make it possible to propose improvements in the future, since the return to face-to-face educational activities will be progressive and virtual courses will continue to be used in university training, including medical education (18). The 30 items of the adapted instrument are oriented to evaluate the perceptions of Peruvian medical students on the quality of virtual courses, which may vary in different contexts and according to the characteristics of the medical schools and the students themselves (19). However, the study evaluated students from most medical schools in Peru and despite potential variations by region, residency and management of each university, the instrument showed adequate reliability in all factors or domains evaluated, in addition to adequate indicators when assessing construct-based validity through exploratory factor analysis.

The first domain of the scale, which seeks to evaluate the students’ perception of the general quality and didactic methodology, groups numerous items; however, in this domain an adequate reliability and factorial loadings were identified. In this sense, the aspects evaluated in this domain are important and consistent with other studies that address issues such as communication between teachers and students and its relationship with the perception of the quality of the teaching methodology (20). Likewise, communication in timely feedback and verification of the understanding of the lessons had a positive impact on the perception of the quality of the course and the didactic methodology (21).

Regarding the second and fifth domain of the scale that evaluates the students’ perception of the design and navigation in the platform or virtual classroom, respectively. Literature was found detailing this domain as an important aspect given that virtual classrooms went from being an optional resource to a unique and necessary environment to continue with university training, thus being their most important characteristics in the evaluation of the quality of virtual courses, due to the fact that students have shown an adequate acceptance towards these platforms or virtual classroom during quarantine periods (22).

The third domain of the scale evaluates the students’ perception of the multimedia resources used in the virtual courses, being a relevant aspect because the resources for conventional learning such as video tutorials and simulators were the most used, probably due to the experience with their use before the COVID-19 pandemic (23). However, the fourth domain of the scale evaluates the academic materials (books, articles, manuals or similar) used in virtual courses that are more frequent in conventional learning and continue to be used because they can be shared virtually; however, in another study a considerable group of students stated that online resources were more useful than the resources used in conventional learning for virtual courses (24).

In our literature review of instruments that evaluate aspects related to virtual courses in the context of COVID-19, we found that they did not address the quality of the courses. Most of them evaluate perceptions about specific aspects of distance education (23-27). This is understandable because the evaluation of a construct such as quality is a challenge because it is a variable concept and even more so because it is used to assess adequate or inadequate quality, in this sense the score of the instrument adapted in this study is referential to the quality of virtual courses. Therefore, this scale represents an opportunity to make evaluations closer to reality with an instrument with adequate sources of validity and reliability in order to make the necessary improvements in virtual courses, based on consultation with students, who play the fundamental role of users of the educational service provided by medical schools and who can give an important perspective on the training they receive or even provide recommendations for the continuous improvement of these same courses.

The study has certain limitations, since it evaluates the virtual courses corresponding to the 2020 academic year that ended on different dates in 2021 for medical schools in Peru, which predisposes to a greater effect of memory bias in some cases, likewise, some students influenced by the social desirability bias could have chosen to favorably describe the courses that were provided by their universities. In addition, there are some particular situations about problems with the virtual courses that could not be evaluated in the instrument but that the students expressed in a blank question placed in each section of the survey and given the variability of these comments we suggest that they could be better addressed with qualitative studies (14). Also, being a virtual survey disseminated through social networks, it has a reach to a specific group of students who share certain characteristics in the use of these platforms (Facebook, Whatsapp and Instagram), which could affect the way in which they evaluate the quality of virtual courses.

However, despite the above, the study also has strengths that lie in the inclusion of students from most medical schools in Peru from different academic years that provide a greater scope of application of the survey, although it was not possible to survey students who performed face-to-face activities in the internship since they had graduated from the university at the time of the survey and because they performed face-to-face activities with a focus more centered on the acquisition of clinical and surgical competencies. This could be addressed by future studies where appropriate instruments are used to assess students’ perceptions of virtual courses and face-to-face activities.

In conclusion, the scale to evaluate the quality of virtual courses developed in the context of COVID-19 for Peruvian medical students shows adequate evidence of validity and reliability, and with its 30 items and five domains, it provides a useful representation of the quality of virtual courses that can help in the development of more accurate evaluations and the consequent continuous improvement of these courses.

## Data Availability

The data produced for this work are contained in the manuscript

## Author contributions

Claudio Intimayta-Escalante (CIE), Rubí Plasencia-Dueñas (RPD) and Ronald Castillo-Blanco (RCB) participated in the conception and design of the study; CIE, RPD, RCB, Kevin Flores-Lovon, Janeth N. Nuñez-Lupaca and Mario Chavez-Hermosilla participated in data collection; while CIE and RCB participated in data management and analysis. All authors participated in the interpretation of the results, the writing and review of manuscript. Finally, all authors of this research approved the final version of the manuscript.

## Acknowledgements

The research team is grateful for the support of Tomás Caycho-Rodríguez, Paolo Wong Chero and Carlos J. Toro-Huamanchumo during the early stages of the study.

